# At-home self-collection of saliva, oropharyngeal swabs and dried blood spots for SARS-CoV-2 diagnosis and serology: post-collection acceptability of specimen collection process and patient confidence in specimens

**DOI:** 10.1101/2020.06.10.20127845

**Authors:** Mariah Valentine-Graves, Eric Hall, Jodie Guest, Elizabeth Adam, Rachel Valencia, Isabel Hardee, Katlin Shinn, Travis Sanchez, Aaron J Siegler, Patrick Sullivan

## Abstract

**Background:** Options to increase the ease of testing for SARS-CoV-2 infection and immune response are needed. Self-collection of diagnostic specimens at home offers an avenue to allow people to test for SARS-CoV-2 infection or immune response without traveling to a clinic or laboratory. Before this study, survey respondents indicated willingness to self-collect specimens for COVID-related tests, but hypothetical willingness can differ from post-collection acceptability after participants collect specimens.

**Methods:** 153 US adults were enrolled in a study of the willingness and feasibility of patients to self-collect three diagnostic specimens (saliva, oropharyngeal swab (OPS) and dried blood spot (DBS) card) while observed by a clinician through a telehealth session. After the specimens were collected, 148 participants participated in a survey about the acceptability of the collection, packing and shipping process, and their confidence in the samples collected for COVID-related laboratory testing.

**Results:** A large majority of participants (>84%) reported that collecting, packing and shipping of saliva, OPS, and DBS specimens were acceptable. Nearly nine in 10 (87%) reported being confident or very confident that the specimens they collected were sufficient for laboratory analysis. There were no differences in acceptability for any specimen type, packing and shipping, or confidence in samples by gender, age, race/ethnicity, or educational level.

**Conclusions:** Self-collection of specimens for SARS-CoV-2 testing and preparing and shipping specimens for analysis were acceptable in a diverse group of US adults. Further refinement of materials and instructions to support self-collection of saliva, OPS and DBS specimens for COVID-related testing is needed.

**Trial registration:** No intervention was tested in this study

## Background

The global SARS-CoV-2 pandemic has resulted in explosive patterns of transmission in most countries [1,2]. Control programs, where they have been successful, have relied largely on a combination of testing people for SARS-CoV-2 infection, quarantine, and social distancing [3]. The United States, which as of this writing comprises 4.25% of the world’s population and accounts for 31% of global diagnoses of COVID-19 disease, has struggled to launch an effective screening program for the virus [4]. After early failures of government-developed testing kits [5], private laboratories were allowed more flexibility to expand testing platforms [6]. However, overall testing capacity remains well below the levels needed to inform decisions about relaxing social distancing programs: public health officials estimate that a US national capacity of at least 1 million tests per week is required [7], and current capacity is less than a quarter of that for PCR testing [8].

There are a number of barriers to expanding testing [9]. Since the beginning of the epidemic, barriers have included supply chain constraints for rigid swabs [10], shortages of personal protective equipment needed for healthcare workers collecting invasive specimens [11], limited transport media for swab-based specimens, and limited laboratory reagents for testing specimens [6]. Further, there is limited willingness of people to travel into a laboratory or clinic to be tested for SARS-CoV-2 infection [12]. Responding to these challenges calls for the development of alternative specimen collection processes that produce high-quality specimens, are acceptable to a broad segment of the population for screening purposes (i.e., not only for diagnostic testing of those suspected of having COVID-19), and minimize the need for personal protective equipment (PPE) for collection.

The iCollect study recruited 159 US adults for a study of the acceptability and sufficiency of at-home self-collected samples for SARS-CoV-2 PCR and immune response testing. We have previously documented the study protocol [9] and demonstrated that specimens collected at home are considered suitable by telehealth clinicians and sufficient by laboratorians [13]. After participants self-collected specimens, we asked them to rate the acceptability of the self-collection process and of packaging and shipping specimens, and we asked participants for recommendations on how to improve the specimen self-collection process. Here, we describe the participant-reported acceptability of self-specimen collection and participant suggestions to improve the self-collection and shipping process.

## Methods

The methods for the iCollect study have been previously described [9]. Briefly, participants were recruited from databases of participants in previous Emory University studies who had agreed to be recontacted for future research studies and from networks of symptomatic and at-risk individuals. Participants were consented online [14] and were mailed a home collection kit that included supplies and instructions to collect three specimens: a saliva specimen for SARS-CoV-2 polymerase chain reaction (PCR) and antibody testing, an oropharyngeal swab (OPS or “throat swab”) for PCR testing, and a dried blood spot (DBS) card for antibody testing.

Participants collected specimens during a telehealth session with a healthcare provider on a HIPAA-compliant video conferencing service. Specimens were returned to the central study laboratory in a provided mailer and tested in the central laboratory. Participants were compensated $40 in Amazon credit for their time. The study was reviewed and approved by our institution’s human subjects research review committee.

Following specimen collection, the clinicians asked participants for suggestions about how to improve the collection instructions or process. Participants were also administered an online survey that collected data on the acceptability of the collection of the three specimens types, the process of packing and shipping the specimens, and how confident the participants were that they had collected the specimens adequately to be accurate as diagnostic specimens.

Participants were also asked three open-ended questions in the online survey to gather qualitative data on suggestions for improvement of the specimen collection instructions. Specifically, they were asked to document suggestions to improve the clarity of instructions to collect or pack each specimen type. We analyzed the data by describing Likert scale scores (1-5, where 1 was very unacceptable and 5 was very acceptable) proportions for acceptability of specimen collection and packing/shipping and medians and interquartile ranges for each acceptability item. We used Kruskal-Wallace tests [15] to describe differences in median acceptability across different demographic groups (i.e., gender, age, race/ethnicity, education). We used a Bonferroni-type correction to set an experiment-wise p alpha of 0.05, considering the 16 comparisons planned in the analysis [16]. We also combined the very acceptable and acceptable responses to make a dichotomous measure of acceptability (i.e., acceptable or very acceptable was categorized as “acceptable”, and all other responses were considered neutral/not acceptable). We used the same process to evaluate data on the confidence of participants that their specimen was suitable for COVID-related testing.

Telehealth observers recorded verbal participant feedback on the process, including participant suggestions of how to improve collection and shipping materials and instructions. Two coders independently read through all participant comments and created themes independently; they then worked together to develop a codebook that was used to independently code the data. A thematic analysis was conducted [17] seeking to assess emergent themes regarding challenges and areas for improvement for the self-collection and return shipping process. The summarized comment themes, examples and frequencies by specimen type are presented in a tabular format.

## Results

Of the 153 participants who self-collected specimens, 148 (97%) completed the post-collection survey for at least one specimen type and provided data on the acceptability of the specimen collection methods and packing/shipping. Most participants were female (59%), non-Hispanic white (70%), and had at least some education beyond high school (97%); median age was 27-49 years (IQR: 27-49). Most surveys were completed within two days of the specimen collection (100, 68%) with 23 on the same day (16%), 46 on the first (31%), 31 on the second (21%) day after the collection. A total of 82 participants provided 135 responses (some containing more than one code or theme) to open-ended survey items with suggestions for improving the specimen collection process. Fifty comments were provided for the DBS collection (34% of those submitting DBS specimens), 53 for saliva collection (35% of those submitting a saliva specimen), and 32 throat swab collection (21% of those submitting a throat swab specimen).

Acceptability of the collection of all three specimen types was high (>84%, Figure 1). Over 90% of participants rated the DBS collection and packaging and mailing processes as very acceptable or acceptable, and 84-86% of participants rated the saliva specimen and throat swab as very acceptable or acceptable. Few participants reported aspects of the collection as unacceptable or very unacceptable: packaging (3%), DBS collection (4%), saliva collection (5%), and throat swab collection (5%). 6%-11% of participants were neutral on acceptability of these components. The median acceptability rating for the DBS specimen was 4 (scale: 1-5, where 5 is very acceptable) and the median acceptability for saliva, throat swab, and packaging and mailing process were 5 (Figure 2). There were no significant differences in the median acceptability of any specimen type of the packaging process by gender, age, race, ethnicity, or education (data not presented).

**Figure 1:**
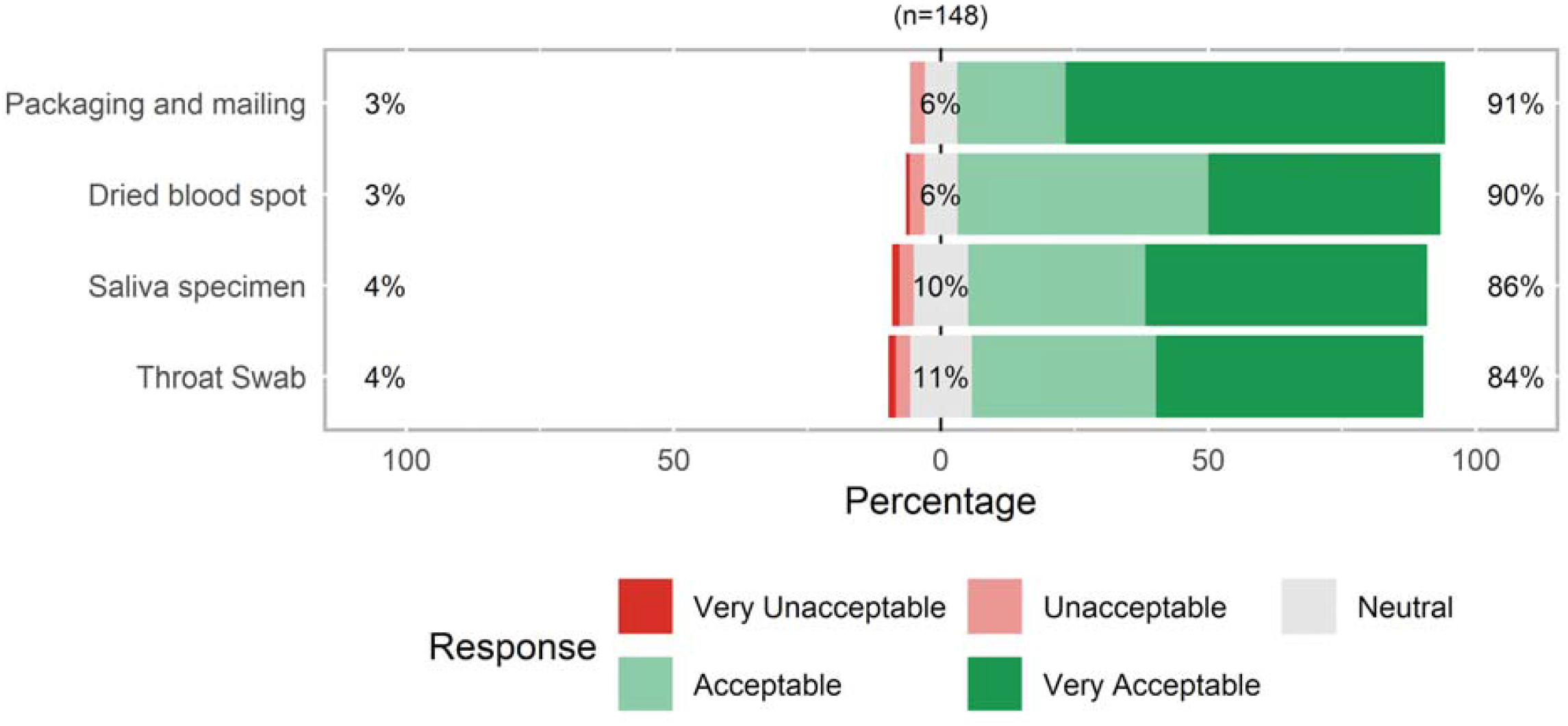
Acceptability of at-home collection, packaging and mailing specimens for SARS-CoV-2 related testing, United States, April 2020.

**Figure 2:**
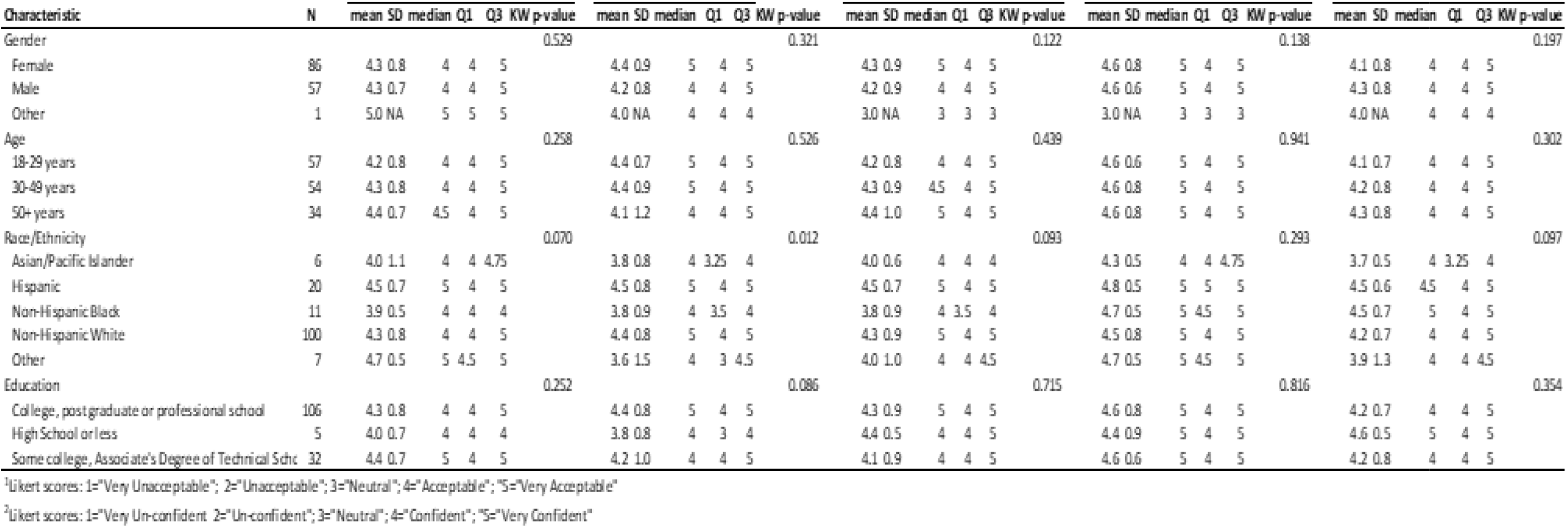
Acceptability of at-home self-collection of saliva, oropharyngeal swab and dried blood spot specimens for SARS-CoV-2-related testing after specimen collection, by gender, age, race/ethnicity, and education, United States, April 2020.

Most participants (87%) reported feeling confident or very confident in the submission of their self-collected specimens for laboratory testing (Figure 3). There were no differences in the median confidence in specimen collection by age, race/ethnicity, or education (data not presented).

**Figure 3:**
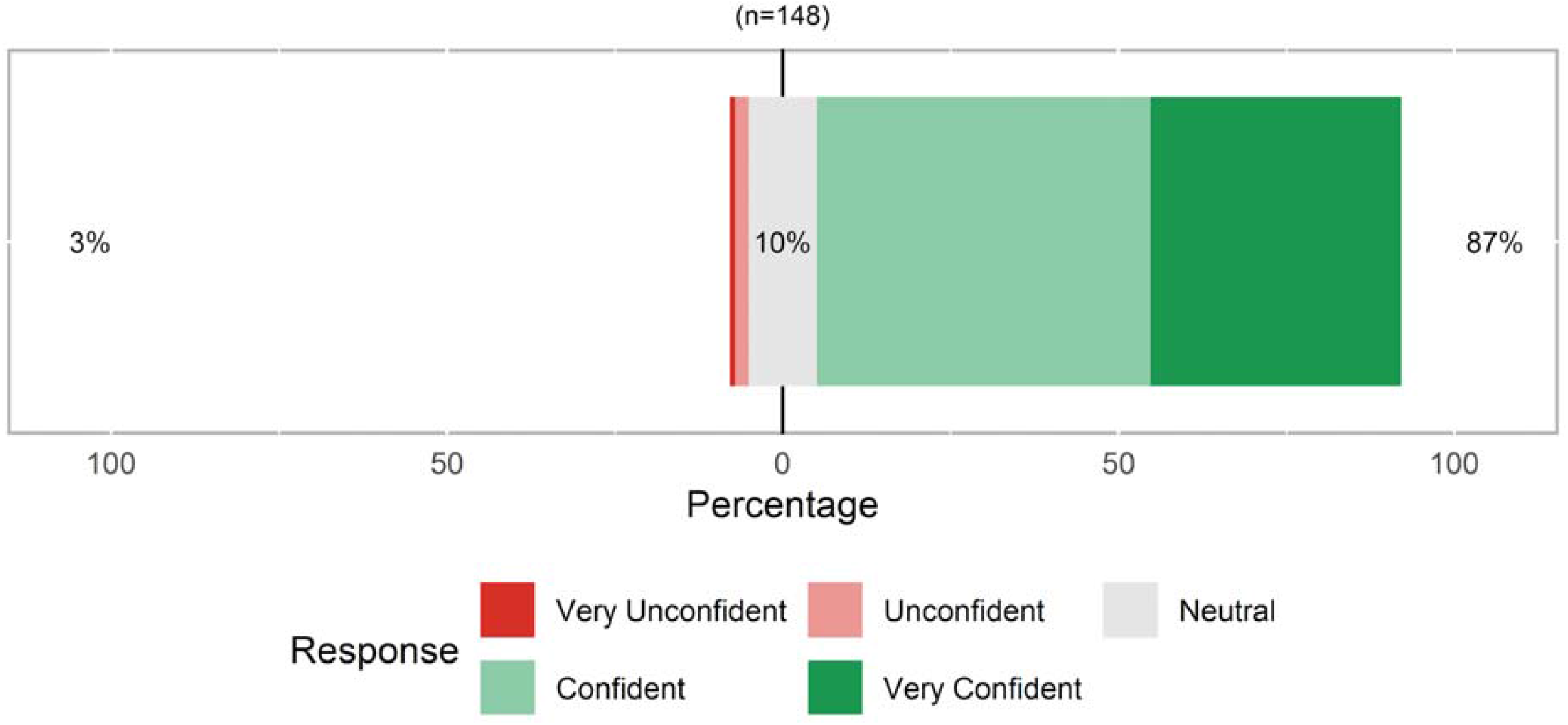
Overall participant confidence in at-home self-collection of saliva, oropharyngeal swab and dried blood spot specimens for SARS-CoV-2-related testing after specimen collection, United States, April 2020.

We identified a number of key themes from participant suggestions, and each was given a code: specimen collection instructions, specimen collection diagrams, kit packaging, device complexity, specimen contamination potential, missing kit materials, and kit shipping (Table 1). Comments categorized as *specimen collection instructions* were most common (n=98 comments) if they described a lack of clarity in the wording or structure of the instruction guide. This included concerns about imprecise or overly technical wording of instructions, inconsistent terminology, lack of sequential ordering of steps, lack of appropriate detail, and inaccessible font type and size. If specifically describing difficulties or suggestions related to the images in the instruction guide, comments were categorized as *specimen collection diagrams*. Thirty comments related to *kit packaging* and detailed difficulties finding the instructions, devices, or supplies inside the kit due to how materials were packaged or organized. Common suggestions within this category were to color code all materials to match the instruction guide, clearly separate and label the three specimen types, and place the instruction guide on top of all other kit materials. Comments in this category were less common (n=20) and were typically about needing more diagrams, more detail with the diagrams and inconsistencies between the images and written text. Twenty comments about *device complexity* detailed difficulties with the collection process due to physical characteristics of the specimen collection devices and offered suggestions to improve clarity of instructions to help participants identify kit components. The saliva device and the DBS collection lancets were described most frequently as being unfamiliar or complex. Comments about *specimen contamination potential* were uncommon (n=9) and related to perceived contamination of the samples during testing due to either the design of the devices or the instructions. In particular, a few participants were concerned with contaminating the OP swab by touching or dropping it before collection. They were also concerned with contaminating the DBS card by touching the paper directly. Four comments related to *missing kit materials* described one or more of the components of the kit missing during testing due to packing error. The participants in this category described missing shipping materials, lancets, or biohazard bags. The final category of kit *shipping* included only two responses related to opportunities to clarify the procedure for packaging and return shipping the kit.

**Table 1.**
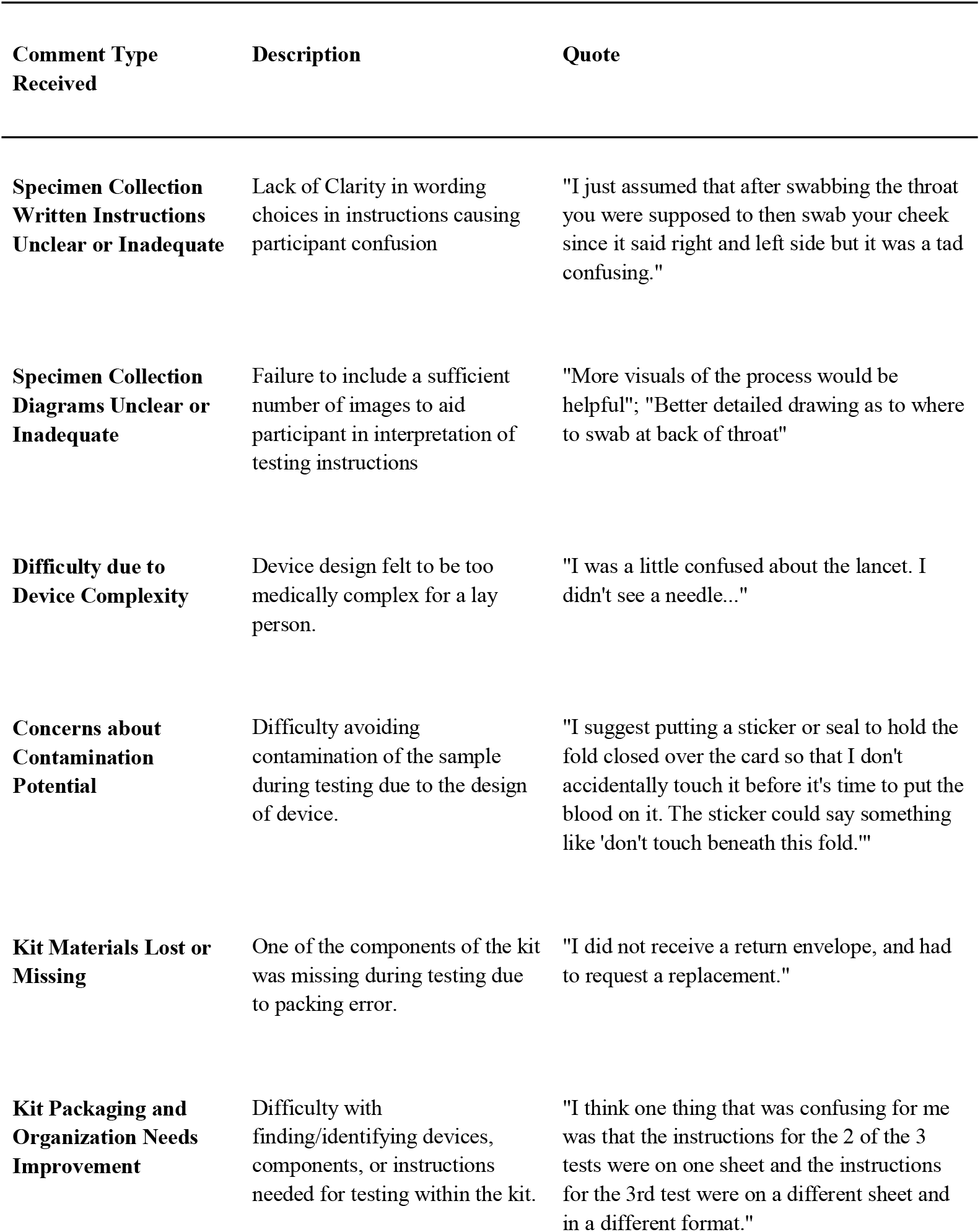
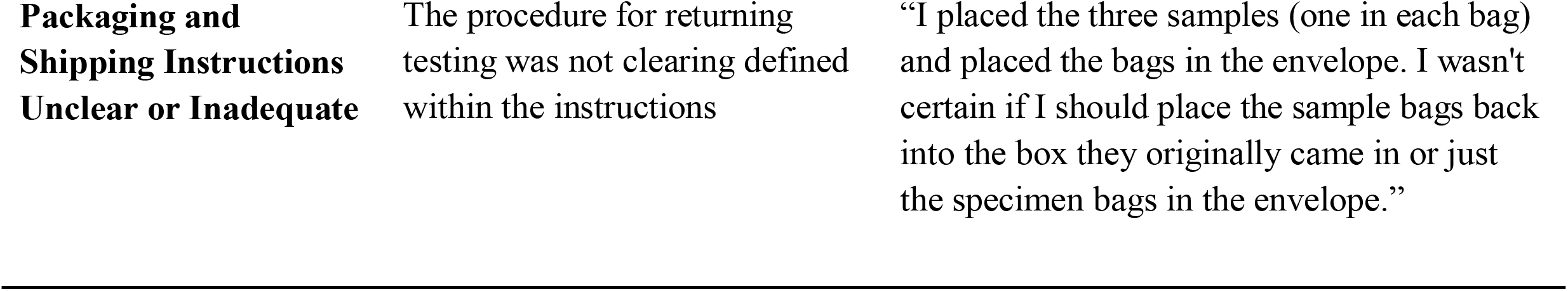
Themes of observations from participants after self-collecting saliva, oropharyngeal swabs and dried blood spot cards for SARS-CoV-2-related testing, United States, April 2020

**Table 2.** Frequency of themes from participants after self-collecting saliva, oropharyngeal swabs and dried blood spot cards for SARS-CoV-2-related testing, United States, April 2020

| **Comment Type Received** | **Frequency of comment type for DBS (n=69)** | **Frequency of comment type for Saliva Collection (n=72)** | **Frequency of comment type for OP Swab (n=42)** |
| --- | --- | --- | --- |
| **Specimen Collection Written Instructions Unclear or Inadequate** | 35 | 42 | 21 |
| **Kit Packaging and Organization Needs Improvement** | 20 | 7 | 3 |
| **Specimen Collection Diagrams Unclear or Inadequate** | 4 | 10 | 6 |
| **Difficulty due to Device Complexity** | 3 | 11 | 6 |
| **Concerns about Contamination Potential** | 5 | 1 | 3 |
| **Kit Materials Lost or Missing** | 1 | 1 | 2 |
| **Packaging and Shipping Instructions Unclear or Inadequate** | 1 | 0 | 1 |

## Discussion

We performed a mixed-methods assessment of the acceptability of three self-collected specimen types for COVID-19 related testing and the confidence of participants in their ability to collect these specimens after they had completed a self-collection process. Our results indicate high acceptability for all three specimen types and high confidence in self-collected specimens. Despite the high overall acceptability of specimen collection, we also identified areas for improving the clarity of instructions and ease of use for devices. These suggestions can be used to further refine the kit and instruction process to improve usability and to decrease potential errors in specimen collection.

We have previously reported that in hypothetical willingness to use at-home specimen collection kits for COVID-19 infection and immune response testing was high (saliva collection: 88% willing; throat swab: 83% willing) among participants in a different, large online sample of US persons. In that study, like the present study, willingness did not vary by race/ethnicity or sex.[18] Younger respondents reported a lower willingness to self-collect an oropharyngeal swab in that study. [18] Further, respondents in the same online study reported high willingness to use home-collection kits to receive COVID-19-related testing for their clinical care (92% reported willingness to use self-collection of saliva and 88% reported willingness to self-collect a throat swab for clinical care).[12] Willingness to use saliva collection and throat swabs for clinical care in that study did not differ by age groups, race/ethnicity, COVID-19 stigma score, COVID-19 knowledge score, or COVID-19 symptomatology. The consistency in the hypothetical willingness to use these methods for research or clinical care and the acceptability of their use after using the kits is notable.

For HIV self-specimen collection systems, hypothetical interest in use was reported to be high before tests became commercially available, but uptake was quite low after they reached the market. [19,20] In the present study, the acceptability of saliva and throat swab collections were higher after participants used them than self-reported hypothetical willingness to use such collection devices in a previous survey. Although a number of factors will eventually influence utilization [21], these data suggest promise for eventual use if these collection methods are made available in the US.

Overall, about half of participants did not provide suggestions to improve the specimen collection process. Of those who did, nearly two-thirds of recommendations were for improvements to the written instructions or diagrams, which can be implemented with minimal redesign of kit components. Our observations of the participants using these kits was the first step in a multi-phased process of developing kits for broader use. We have used this staged process in the past to develop home-use kits for collection of DBS specimens and for home testing devices.[22,23] The data collected from these participants who used a pre-pilot version of the kits will be used to refine instructions. In the case of our PrEP@Home project, we also developed a video (https://vimeo.com/138977095) to illustrate the key steps of the process and to draw attention to potential errors in self-collection that were observed in the pilot phase. In this case, we will use the data from participant suggestions to clarify instructions, redesign the user experience in terms of the assembly and order of kit components, take steps to minimize touching of the DBS card before specimen collection, and include more explicit instructions for packaging and returning the kit. With these modifications in place, we will re-pilot the usability of the modified kit to test the impact of the modifications.

The feedback of the participants is interpreted in light of the evaluation of the specimen quality by clinicians and laboratorians.[24] The specimens collected by these same participants were judged by clinicians to be suitable for laboratory testing (saliva: 97%; throat swab: 96%) and were found to have adequate nucleic acid for RNA PCR diagnostic testing (saliva: 100%; throat swab: 99%).[24]. Comparing the three measures in terms of the assessment of sufficiency of specimen collection, participants had the lowest confidence in the adequacy of their specimen collections; healthcare providers observing the collection and laboratorians processing the specimens were more confident in the sufficiency of the specimens.

Our study has important limitations. Our participants represent a convenience sample, and it is likely that they are a biased sample and may over-represent people interested in COVID-19-related testing generally, or people concerned about their own SARS-CoV-2 infection status. It is unclear how these biases might impact the results of our self-evaluation of acceptability of specimen collection. Further, the lay opinions of participants about their confidence in the sufficiency of specimens they collected may be naive since they were not selected for healthcare or laboratory expertise. However, the results of clinician observation and laboratorian assessment supported their views of the sufficiency of their samples; the triangulation of these three assessments lends credence to the potential sufficiency of self-collected specimens for COVID testing.[25]

The United States is in dire need of options to increase screening for SARS-CoV-2 infection and immune experience, and survey data suggest that people might be more willing to collect specimens at home for research and clinical purposes than to travel into labs or clinics to provide specimens.[12,26] At-home self collection offers possibilities to reduce the exposure of people in need of screening to others by not requiring travel to test; to reduce the need for PPE for invasive screening tests; to offer options for screening of populations without symptoms who might be unwilling to travel to a clinic for testing. Next steps in this development process include modification to kit instructions and assembly, addition of instructions for shipping, development of video instructions (emphasizing areas of lack of clarity or participant concern), and reassessment of kit usability and performance with another group of participants observed by a telehealth clinician in an intended use environment (e.g., home collection).

## Data Availability

Data will be available upon request to the authors

## List of abbreviations

DBS: Dried blood spot
OPS: Oropharyngeal swab
PCR: Polymerase chain reaction
PPE: Personal protective equipment

## Acknowledgements

This work was supported by the National Institutes of Health (3R01AI143875-02S1). The authors acknowledge Sarah Johnson and Iaah Lucas for their support of the research process.

